# Child protection contact among children of culturally and linguistically diverse backgrounds: a South Australian linked data study

**DOI:** 10.1101/2022.03.18.22272447

**Authors:** R. Abdul Rahim, R. Pilkington, A. M. Procter, A. Montgomerie, M. N. Mittinty, K. D’Onise, J. Lynch

## Abstract

**Aim:** To determine the cumulative incidence of child protection (CP) system contact, maltreatment type, source of reports up to age 7, and sociodemographic characteristics for Culturally and Linguistically Diverse (CALD) Australian children.

**Methods:** A South Australian (SA) whole-of-population linked administrative data study of children followed from birth up to age 7, using child protection, education, health, and birth registrations data. Participants: SA born children enrolled in their first year of school from 2009-2015 (*N* = 76 563). CALD defined as non-Aboriginal or Torres Strait Islander, spoken language other than English, Indigenous or Sign, or had at least one parent born in a non-English speaking country. Outcomes measures: For CALD and non-CALD children the cumulative incidence of CP reports up to age 7, relative risk and risk differences for all CP contact (reporting through to out-of-home care (OOHC)) and age, primary maltreatment type, reporter type, and socioeconomic characteristics were estimated. Sensitivity analyses explored population selection and different CALD definitions.

**Results:** By age 7, 11.2% of CALD children were screened in compared to 18.8% of non-CALD (RD 7.6 percentage points (95% CI: 6.9-8.3)), and 0.6% of CALD children experienced OOHC compared to 2.2% of non-CALD (RD 1.6 percentage points (95% CI: 1.3-1.8)). Among both groups, most common abuse type was emotional and the most common reporter types were police and education sector. Socioeconomic characteristics were broadly similar. Sensitivity analyses results were consistent with primary analyses.

**Conclusion:** By age 7, contact with any level of child protection was lower for CALD compared to non-CALD children. Estimates based on primary and sensitivity analyses suggested CALD children were 5 to 9 percentage points less likely have a report screened-in, and from 1.0 to 1.7 percentage points less likely to have experienced OOHC.

**What is already known on this topic:** In Australia, Aboriginal and/or Torres Strait Islander children are over represented in the child protection system.

In England and United States child protection contact vary between ethnicities with over-representation of Black and African American, and under-representation of Asian children.

Disparities in child protection contact between ethnicities are largely driven by socioeconomic disadvantage.

**What this paper adds:** CALD children were less likely to have contact across all levels of child protection from reporting to OOHC. By age 7, 11.2% of CALD children were screened-in compared to 18.8% of non-CALD.

Sensitivity analysis suggested CALD children were 5 to 9 percentage points less likely to have a report screened-in, and from 1.0 to 1.7 percentage points less likely to have experienced OOHC.

CALD and non-CALD groups did not markedly differ by the type of maltreatment, source of report, or on background socioeconomic factors.

## Introduction

Child maltreatment is abuse and/or neglect of a child, and has long been recognised as an important public health issue.^1-3^ In 2019-20 child protection (CP) agencies in Australia received 486,300 notifications for 275 000 children.^4^ In South Australia (SA), 25% of children were notified at least once to CP services by the age of 10 years.^5^ Adverse effects of maltreatment on mental^6, 7^ and physical health, education attainment, labour market participation,^8^ and increased risk of antisocial behaviours^9 10^ have been shown in previous research.

Racial or ethnic disparities in CP contact are well recognised.^11^ In the United States,^12-14^ Canada,^15^ Australia^4, 16^ and New Zealand^17^ children of First Nations communities are over-represented in the CP system, as a consequence of colonialism, dispossession and systemic discrimination. The CP experience is heterogeneous within subgroups of racialized or ethnic minority groups. In England, the rate of children substantiated was higher in Mixed heritage (84 per 10 000) compared to White (50 per 10 000) children.^18^ Asian children had a lower rate compared to White at 23 per 10 000 children.^18^ However, the rate of Black (87 per 10 000) and Mixed heritage (99 per 10 000) children in OOHC far exceeded that of White children (64 per 10 000).^18^ In the United States approximately 20% of African American children and between 10 to 13% of Latino/Hispanic children were substantiated for maltreatment compared to 10% of White children.^12, 13^ Conversely the cumulative incidence of substantiated maltreatment in Asian and Pacific Islander children was less than 5%.^12, 13^ In both countries the disparities in contact patterns between White and racialized or ethnic minority children are largely driven by socioeconomic disadvantage.^11-13, 18-23^ Little is known about the CP contact of children from non-Indigenous ethnic minority groups in Australia.

Being from a “Culturally and Linguistically Diverse” (CALD) background is a term unique to Australia. It refers to people who are not of the mainstream English-speaking Anglo-Celtic group and do not identify as Aboriginal or Torres Strait Islander (Australia’s First Nations population). Although CALD is widely used in public and policy discourse, there are no consensus guidelines on enumerating this heterogeneous population.^24^ Despite the availability of data collection frameworks for CALD characteristics such as country of birth and language spoken, the concept of CALD lacks an operational definition.^24, 25^ Applying various definitions to data from the 2016 Australian Census, we showed the estimated proportion of CALD children aged 0-17 ranged from 11% to 44%.^25^

Only four previous studies have examined CP contact for CALD children in Australia but none were able to provide incidence or prevalence estimates due to lack of population denominators.^26-29^. Our objectives are descriptive,^30, 31^ so we report a primary analysis of the cumulative incidence of child maltreatment, maltreatment type, and source of notification up to age 7, as well as sociodemographic characteristics using our preferred definition to identify CALD and non-CALD children attending government schools in South Australia (SA). Sensitivity analyses assessed robustness of estimates to study population selection and CALD definition, and the over-representation of Aboriginal and/or Torres Strait Islander children in the CP system. This study utilised administrative data of all reports to the statutory child protection agency. These reports are a marker of maltreatment, of which the true burden in the community is unknown.

## Methods

### Data sources

This study used whole population de-identified data from the SA Better Evidence, Better Outcomes Linked Data (BEBOLD) platform. Child protection records were linked with data from the Perinatal Statistics Collection, Births, Deaths and Marriages Registry, Government School Enrolment Census and the 2009, 2012 and 2015 Australian Early Development Census (AEDC). Linkage was conducted by SANT Datalink, an independent agency.^32^ The estimated false positive linkage error is 0.4%.^33^

### Population

Primary analysis included children born in SA who started in a government school between 2009 and 2015 (N=76 563), corresponding to children born 2003-2010. Additional analysis describing socioeconomic characteristics was limited to children with complete information recorded at birth (n=72 209) (Figure1).

**Figure 1:**
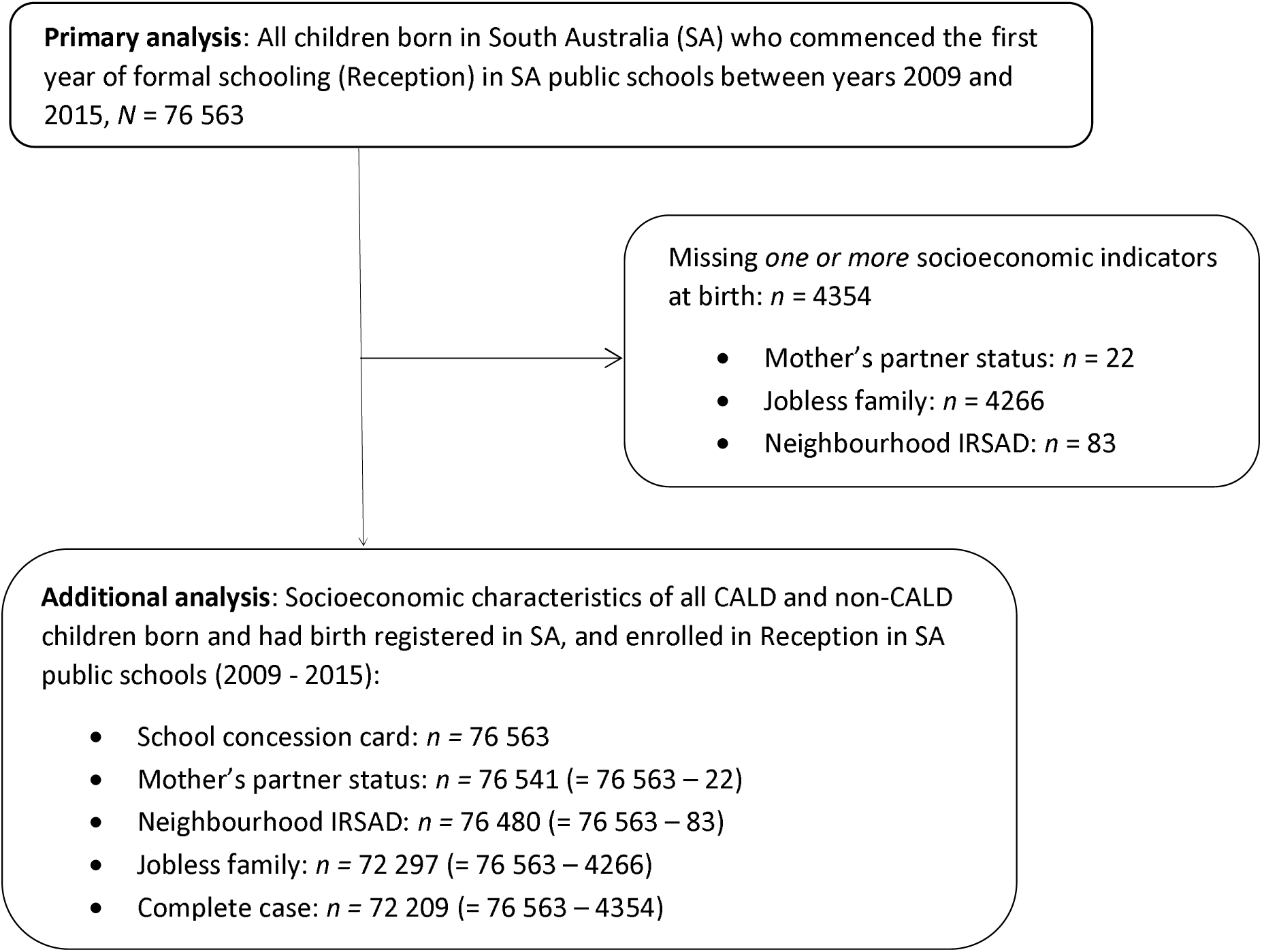
Flowchart of study population and sample for primary and additional analysis

### Child protection contact

CP data cover the entire child protection system from initial reports by mandatory and non-mandatory sources to experience of out-of-home-care (OOHC). These include initial reports, reports not screened-in, screened-in reports not investigated, investigations not substantiated, substantiations, and OOHC. Other CP data included child age, reporter type (e.g. health, education, police), alleged (screened-in reports) and substantiated maltreatment type.

### CALD identification

Our preferred indicator of CALD was based on a combination of language(s) spoken at home and parental country of birth. These items are only available in the School Enrolment Census (SEC) for children attending government schools. As many CALD children born in Australia have parents who were immigrants, the child’s country of birth was not used. We defined CALD children as non-Aboriginal or Torres Strait Islander who spoke a first or second language other than English, Celtic, Australian Indigenous or sign at home, and/or had at least one parent born in countries other than Australia, New Zealand, UK, Ireland, USA, and Canada. Non-CALD were all Aboriginal or Torres Strait Islander and non-Indigenous peoples who spoke English, Celtic, any Australian Indigenous or sign language at home and had both parents born in Australia, New Zealand, UK, Ireland, USA, Canada or South Africa.

For sensitivity analysis to include children in non-government schools, we used data from the AEDC to define CALD. This is similar to the definition above except that the child’s country of birth was used, as parental country of birth was not available. Those from a non-English language background and/or born in non-English speaking countries were classified as CALD.

### Socioeconomic disadvantage

Socioeconomic indicators were obtained from perinatal and birth registration data, and SEC. This included mother’s partner status, mother’s residential postcode, and father’s and mother’s workforce participation. Jobless families were those with no parent(s) participating in the workforce.^34^ The Index of Relative Socioeconomic Advantage and Disadvantage (IRSAD) was based on residential postcode, and a school concession card that subsidises education expenses.^35^

## Analysis

### Primary analysis

Cumulative incidence of all levels of contact with child protection up to age 7 years was calculated by dividing the number of CALD and non-CALD children within each level of contact by the total number of children. We estimated the prevalence of ever having child protection contact by age 4 (to capture experiences before school), and by age 7, and generated relative risks and risk differences. Within the population of children who were reported we 1) calculated proportions of children by age at first notification in infancy (<1 year), early childhood (1 to 4 years) and the first three years of formal schooling (5 to 7 years), by sex; 2) calculated reporter type for reports up to age 4, and between ages 5 to 7; and 3) examined primary alleged and substantiated maltreatment type for all reports up to age 7.

### Additional analysis

We explored socioeconomic characteristics of families of CALD and non-CALD children on a subset of the primary analysis population with complete data.

### Sensitivity analysis

#### Agreement between two measures of CALD

We calculated classification agreement in a sub-population with CALD indicators based on the SEC and AEDC. This population comprised children with an AEDC in government schools in 2009, 2012 or 2015, and included children not born in SA. To account for imbalanced outcomes we used the Agreement Coefficient (AC_1_) as a measure of concordance^36, 37^ between the measures of CALD. (See Supplementary Table S1)

#### Variations to the study population

We undertook four sensitivity analyses. 1) We included children attending independent schools in addition to those in government schools, and used an alternative definition of CALD derived from the AEDC (where child’s country of birth was used as parental country of birth was not available). 2) We limited the analysis to an overlapping subset of the study population comprised of children who sat the AEDC in a government school in 2009, 2012 and 2015, and who were identified as CALD by either definition. 3) Given the over-representation of Aboriginal and Torres Strait Islander children in the child protection system, the third sensitivity analysis excluded those children from the non-CALD comparison group. 4) We repeated the primary, and the first and second sensitivity analyses to include all children, including migrant children who were not born in SA and so would not have a perinatal or birth record but could have AEDC and school records.

### Ethics approval

Ethics approval was granted by the Human Research Ethics Committee of the South Australian Department of Health and Ageing (SA Health) (ref HREC/13/SAH/106; HREC/13/SAH/106/AM18), the University of Adelaide Human Research Ethics Committee (H-185-2011), and the Aboriginal Health Council Research Ethics Committee (ref 04-13-538).

## Results

### Primary analysis

Of the 76 563 children in the study population, 12.1% were CALD. Figure 2 shows the cumulative incidence of at least one report was lower in CALD children at all ages. The cumulative incidence of CP reports was 3.6% and 7.5% at under age 1 for CALD and non-CALD children respectively. By age 7 the cumulative incidence of CP reports was 21.4% for CALD and 35.1% for non-CALD children. This corresponds to risk differences of 3.9 percentage points at age 0, and 13.7 percentage points at age 7. Cumulative incidence up to age 7 by different birth cohorts is shown in Figure S1.

**Figure 2:**
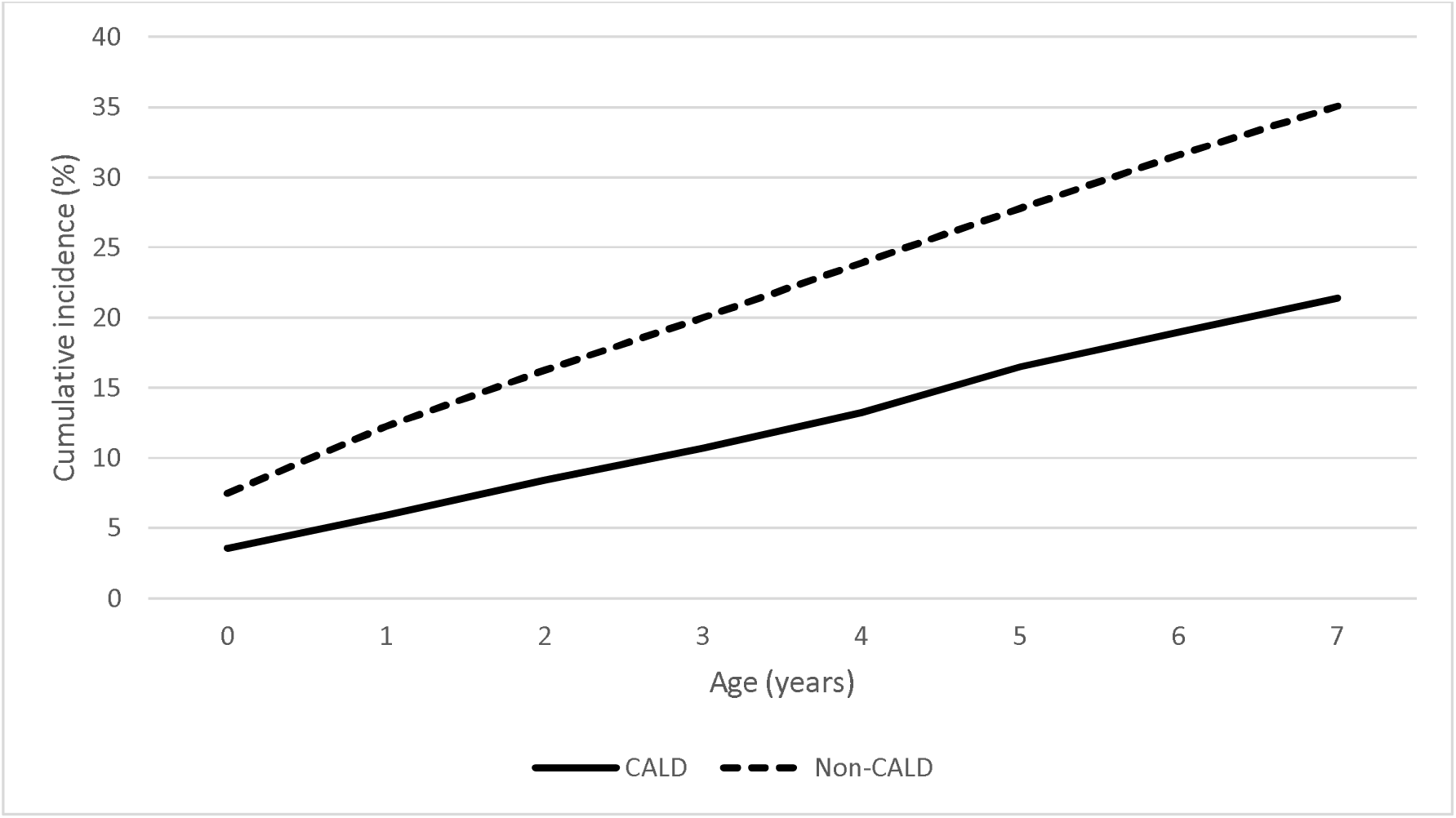
Cumulative incidence of child protection reports up to age 7 and enrolled in school from 2009-2015, followed to the end of 2017 (n= 76 563)

Table 1 shows all CP contact patterns. By ages 4 and 7, the risk of child protection contact was lower for CALD compared to non-CALD children across all levels of contact from reports to OOHC. For example, by age 7, the risk difference between CALD and non-CALD children by age 7 for screened-in was 7.6 percentage points, whilst for OOHC it was 1.6 percentage points. Risk ratios increased with greater intensity of contact so that by age 7 non-CALD children had over three times the risk of being in OOHC (RR = 3.52, 95% CI 2.70 – 4.58).

**Table 1:**
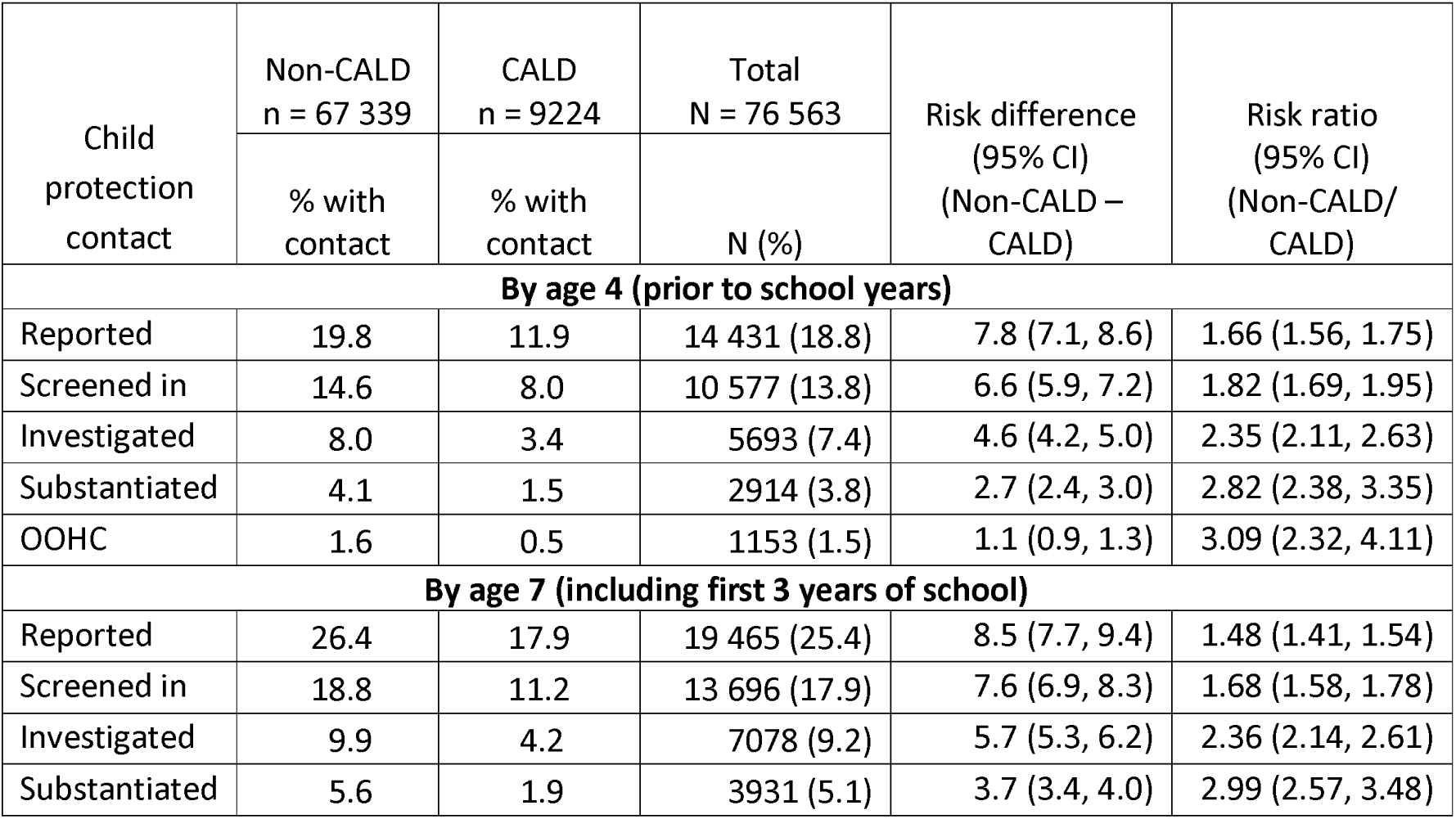
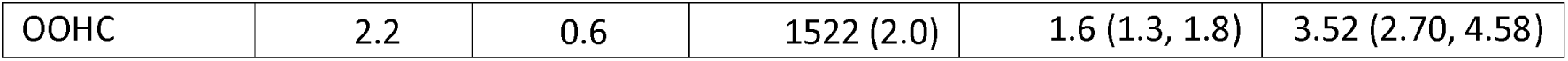
Child protection contact by ages 4 and 7 years for children born in SA and in SA public schools between 2009 and 2015, followed to the end of 2017 (n=76 563)

| Child protection contact | Non-CALD<br>n = 67 339 | CALD<br>n = 9224 | Total<br>N = 76 563 | Risk difference<br>(95% CI)<br>(Non-CALD – CALD) | Risk ratio<br>(95% CI)<br>(Non-CALD/<br>CALD) |
| --- | --- | --- | --- | --- | --- |
|  | % with contact | % with contact | N (%) |  |  |
| By age 4 (prior to school years) |  |  |  |  |  |
| Reported | 19.8 | 11.9 | 14 431 (18.8) | 7.8 (7.1, 8.6) | 1.66 (1.56, 1.75) |
| Screened in | 14.6 | 8.0 | 10 577 (13.8) | 6.6 (5.9, 7.2) | 1.82 (1.69, 1.95) |
| Investigated | 8.0 | 3.4 | 5693 (7.4) | 4.6 (4.2, 5.0) | 2.35 (2.11, 2.63) |
| Substantiated | 4.1 | 1.5 | 2914 (3.8) | 2.7 (2.4, 3.0) | 2.82 (2.38, 3.35) |
| OOHC | 1.6 | 0.5 | 1153 (1.5) | 1.1 (0.9, 1.3) | 3.09 (2.32, 4.11) |
| By age 7 (including first 3 years of school) |  |  |  |  |  |
| Reported | 26.4 | 17.9 | 19 465 (25.4) | 8.5 (7.7, 9.4) | 1.48 (1.41, 1.54) |
| Screened in | 18.8 | 11.2 | 13 696 (17.9) | 7.6 (6.9, 8.3) | 1.68 (1.58, 1.78) |
| Investigated | 9.9 | 4.2 | 7078 (9.2) | 5.7 (5.3, 6.2) | 2.36 (2.14, 2.61) |
| Substantiated | 5.6 | 1.9 | 3931 (5.1) | 3.7 (3.4, 4.0) | 2.99 (2.57, 3.48) |
| OOHC | 2.2 | 0.6 | 1522 (2.0) | 1.6 (1.3, 1.8) | 3.52 (2.70, 4.58) |

Table 2 shows alleged and substantiated maltreatment type for screened-in reports. Emotional abuse was the most common alleged maltreatment, affecting around 60% of CALD and non-CALD children by age 7. Half of CALD and non-CALD children were substantiated for emotional abuse. CALD children were less likely to be screened in or substantiated for neglect and sexual abuse compared to non-CALD children. About 40% of CALD children screened in were allegedly physically abused. This reduced to less than a third of CALD children substantiated. The proportion of CALD children who were allegedly sexually abused was approximately half that of non-CALD children.

**Table 2:** Primary alleged (n=13 696) and substantiated (n=3931) maltreatment type for all notifications up to age 7 years†, followed to the end of year 2017

| Maltreatment type | Alleged (n=13 696) |  |  |  | Substantiated (n=3931) |  |  |  |
| --- | --- | --- | --- | --- | --- | --- | --- | --- |
|  | Non-CALD |  | CALD |  | Non-CALD |  | CALD |  |
|  | n | % | n | % | n | % | n | % |
| Emotional abuse | 7970 | 62.9 | 616 | 59.7 | 1848 | 49.2 | 85 | 49.4 |
| Neglect | 7219 | 57.0 | 352 | 34.1 | 2029 | 54.0 | 61 | 35.5 |
| Physical abuse | 5632 | 44.5 | 410 | 39.7 | 859 | 22.9 | 49 | 28.5 |
| Sexual abuse | 3477 | 27.5 | 166 | 16.1 | 406 | 10.8 | 8 | 4.7 |

† children can have more than one type of maltreatment due to multiple screened-in or
substantiated reports

Age patterns showed that of children reported to child protection, 19.9% of CALD compared to 28.3% of non-CALD children were reported before age 1, and 33.4% of CALD children were reported when school aged (5 to 7 years), compared with 25.2% of non-CALD children. There were few sex differences with the exception of a higher proportion of CALD children substantiated by age 7 were females (55.2%) (age and sex data not shown).

Figure 3A shows the distribution of reporter type was similar with police as the main source under age 4, followed by reports from health, and family and personal social networks. Figure 3B shows that schools and police accounted for over half of reports in CALD and non CALD children aged 5 to 7 years.

**Figure 3A:**
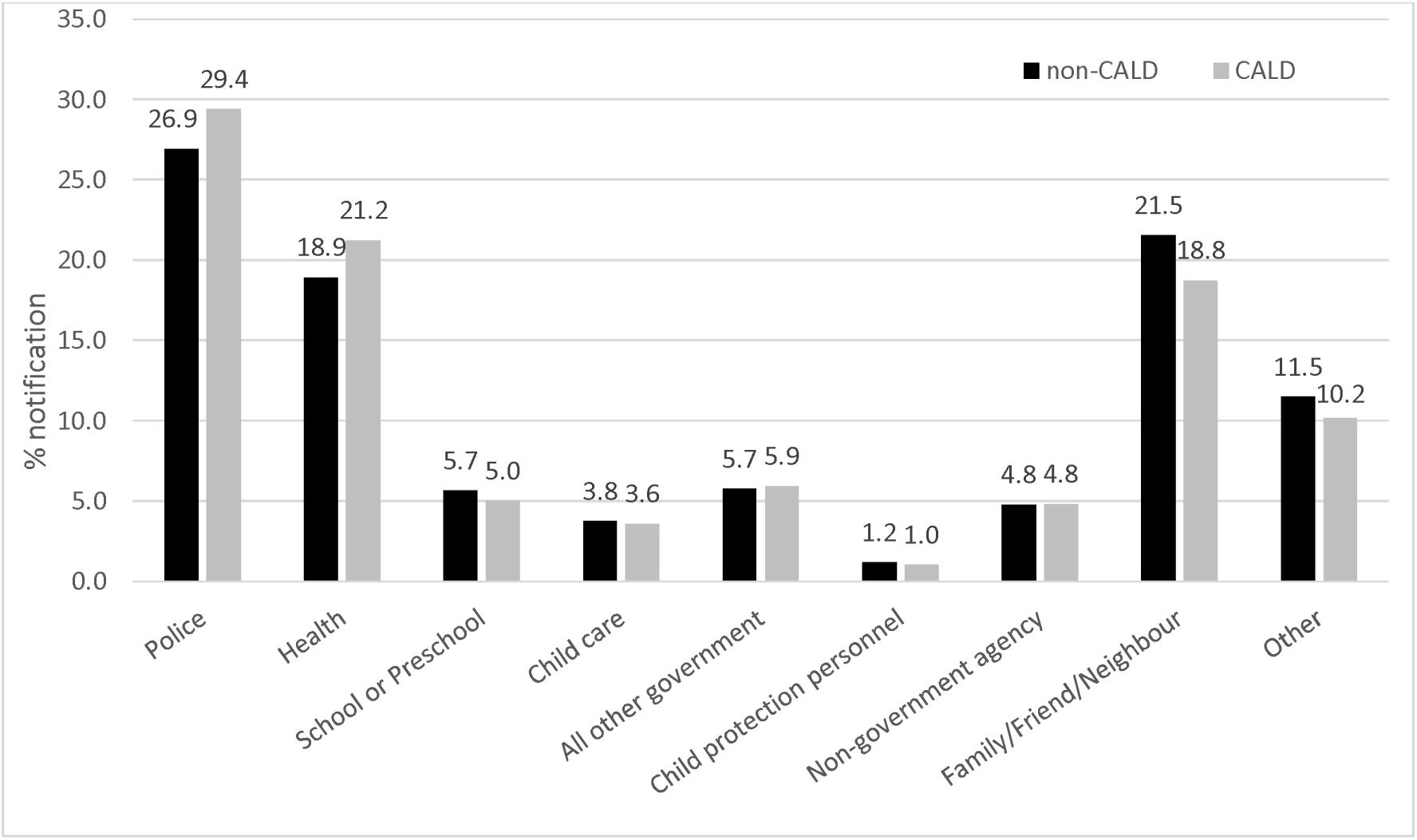
Sources of all reports for children first reported by age 4 years. Denominator: number of reports for children born in SA and attended reception in SA public schools between years 2009 and 2015 (*n* = 50 790). Each child may be reported more than once.

**Figure 3B.**
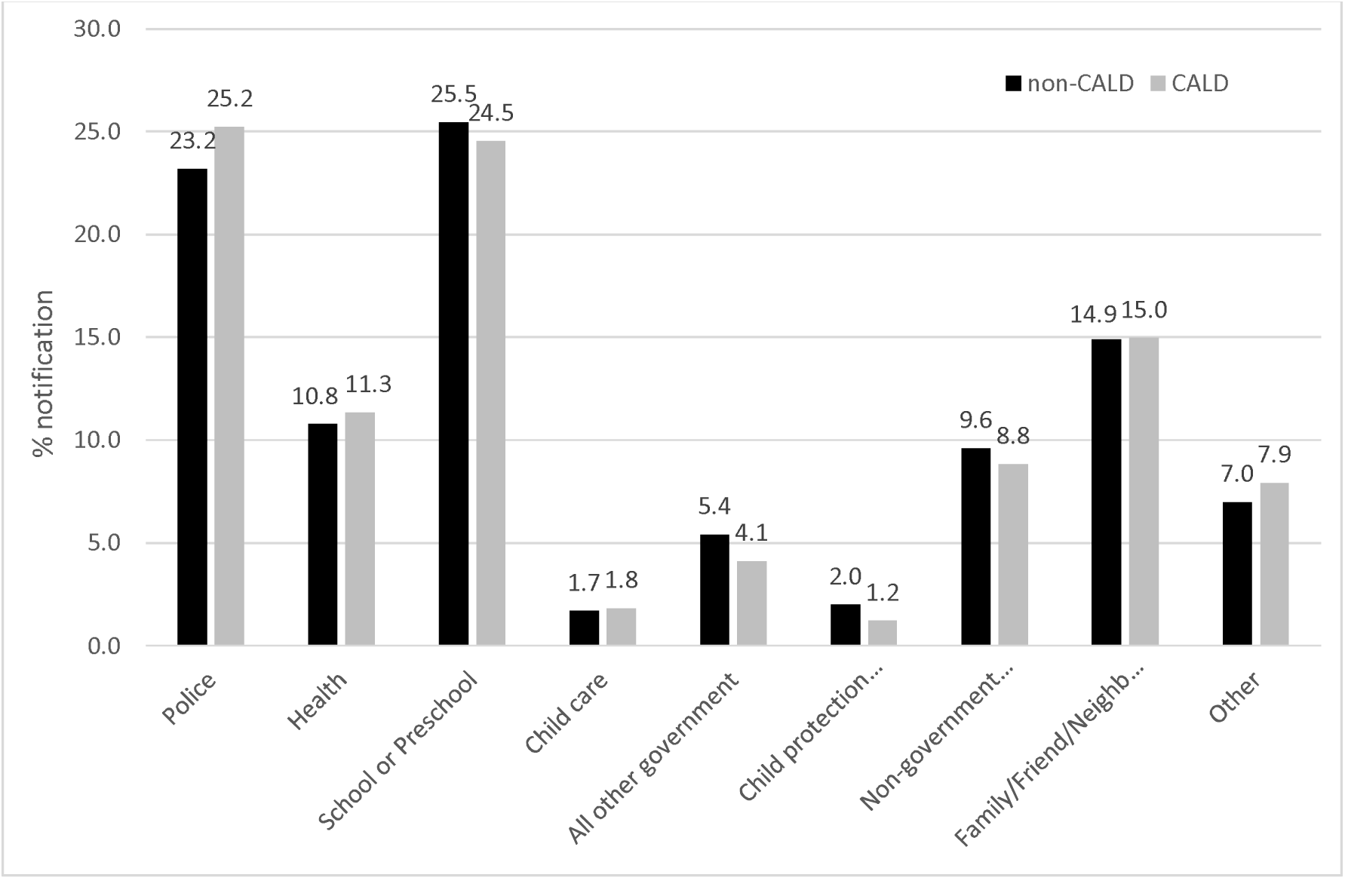
: Sources of all reports for children first reported between ages 5 and 7 years. Denominator: number of reports for children born in SA and attended reception in SA public schools between years 2009 and 2015 (*n* = 40 968). Each child may be reported more than once.

### Socioeconomic characteristics

Table 3 shows few socioeconomic differences, except mothers of CALD children were more likely to be partnered at birth.

**Table 3:** Socioeconomic characteristics of CALD and non-CALD children. Study population: complete cases of children born and had birth registered in SA who attend reception in a SA public school between years 2009 and 2015, followed up until the end of year 2017 (*n* = 72 209)

|  | Non-CALD<br>N = 63 771 | CALD<br>N = 8438 | Total<br>N = 72 209 |
| --- | --- | --- | --- |
|  | Col % | Col % | N (Col %) |
| Mother's partner status |  |  |  |
| Partner | 84.3 | 90.2 | 85.0 |
| No partner | 15.7 | 9.8 | 15.0 |
| Jobless family |  |  |  |
| No | 89.6 | 86.1 | 89.2 |
| Yes | 10.4 | 13.9 | 10.8 |
| IRSAD area-based decile within Australia based on IRSAD for child year of birth |  |  |  |
| Most disadvantaged | 17.7 | 16.3 | 17.5 |
| 2 | 10.0 | 10.2 | 10.0 |
| 3 | 11.3 | 8.9 | 11.0 |
| 4 | 13.7 | 12.2 | 13.5 |
| 5 | 11.2 | 10.6 | 11.1 |
| 6 | 6.3 | 6.8 | 6.3 |
| 7 | 8.1 | 9.9 | 8.3 |
| 8 | 10.2 | 10.5 | 10.3 |
| 9 | 8.4 | 10.8 | 8.7 |
| Most advantaged | 3.1 | 3.8 | 3.2 |
| School concession card |  |  |  |
| No | 80.7 | 80.6 | 80.7 |
| Yes | 19.3 | 19.4 | 19.3 |

### Sensitivity analyses

Agreement between the two definitions of CALD (AC_1_=0.92) was high (see Table S1).

Sensitivity analyses were consistent with the primary analysis where the risk of all forms of contact was lower in CALD children. Primary analysis showed that by age 7, the risk difference between CALD and non-CALD was 7.6 percentage points. Sensitivity analyses bounded these risk differences from 5 to 9 percentage points. For OOHC risk differences ranged between 1.0 and 1.7 percentage points. Results of the sensitivity analyses are shown in Tables S2-S7.

## Discussion

Using whole-of-population linked administrative data we demonstrated that the risk of all forms of contact with the CP system up to age 7 was lower for CALD than non-CALD children. Our primary analysis showed that by age 7, CALD children were 7.6 percentage points (95% CI: 6.9-8.3) less likely to have been screened-in, and of 1.6 percentage points (95% CI: 1.3-1.8) less likely to have experienced OOHC than non-CALD children. In the absence of self-identification, there is no gold standard for defining CALD populations. Using different definitions of CALD applied in different, and the same populations showed consistent findings. The magnitude of risk differences depends on which definitions and populations are used, but our estimates show that CALD children were from 5 to 9 percentage points less likely to have reached a risk threshold to be screened-in, and from 1.0 to 1.7 percentage points less likely to have experienced OOHC. In our study, CALD and non-CALD families appeared to have broadly similar socioeconomic profiles.

Many CALD people would identify as immigrants, but we were not able to examine this because data on immigrant status is not currently available from the Australian government. Information on immigration status may enable further analysis to identify high need CALD subpopulations, such as refugees who are identified by visa subclass.

As this is the first study to attempt to describe the child protection experience of CALD children and families, we did not do a further breakdown of CALD subgroups, despite the obvious heterogeneity in a diverse CALD population. Creating broad categories (e.g. Asian, African) as has been done in studies elsewhere ^18, 19^ may not support meaningful interpretation, and may inadvertently stigmatise subsets of the CALD community. Also, it raises the risk of re-identification given the small numbers that would be generated if CALD children were to be meaningfully categorised according to language spoken or parental country of birth.

## Conclusion

Up to age 7, CALD children had lower risk of contact across all levels of contact with the CP system in South Australia, from notification to OOHC placement. CALD and non-CALD groups did not markedly differ by the type of maltreatment, source of notification, or on background socioeconomic factors. This general pattern of lower risk among CALD populations was consistent across sensitivity analyses exploring different definitions of CALD and using different study populations.

## Data Availability

Data produced in this study is not available

## Acknowledgments

JL was awarded a National Health and Medical Research Council (NHMRC) Australia Fellowship (570120); the NHMRC Centre of Research Excellence (1099422); and the NHMRC Partnership Project Grant (1056888); RMP, AM, was supported by funds from the NHMRC Centre for Research Excellence; AMP was supported by funds from the Westpac Scholars Trust; and RAR was supported by the University of Adelaide Faculty of Health and Medical Sciences Divisional Scholarship.

## References

1. Gilbert R, Spatz Widom C, Browne K, Fergusson D, Webb E, Janson S. Burden and consequences of child maltreatment in high-income countries. Lancet. 2009;373(9657):68–81.

2. Malvaso C, Pilkington R, Montgomerie A, Delfabbro P, Lynch J. A public health approach to preventing child maltreatment: An intelligent information infrastructure to help us know what works. Child Abuse & Neglect. 2020;106(104466).

3. Higgins D, Lonne B, Herrenkohl TI, Scott D. The Successes and Limitations of Contemporary Approaches to Child Protection. In: Lonne B, Higgins D, Scott D, Herrenkohl TI, editors. Re-Visioning Public Health Approaches for Protecting Children. Child Maltreatment, Contemporary Issues in Research and Policy, 2211-9701;9. 1st ed. Switzerland: Cham: Springer International Publishing; 2019.

4. Australian Institute of Health and Welfare. Child protection Australia 2019-20. Canberra: AIHW, 2021 no.74. Cat. no. CWS 78.

5. Pilkington R, Grant J, Chittleborough C, Gialamas A, Montgomerie A, Lynch J. Child Protection in South Australia. The University of Adelaide, BetterStart Child Health and Development Research Group School of Public Health; 2017.

6. Croft J, Heron J, Teufel C, Cannon M, Wolke D, Thompson A, et al. Association of Trauma Type, Age of Exposure, and Frequency in Childhood and Adolescence With Psychotic Experiences in Early Adulthood. JAMA Psychiatry. 2019;76(1):79–86.

7. Mills R, Scott J, Alati R, O’Callaghan M, Najman JM, Strathearn L. Child maltreatment and adolescent mental health problems in a large birth cohort. Child Abuse & Neglect. 2013;37(5):292–302.

8. Mills R, Kisely S, Alati R, Strathearn L, Najman JM. Cognitive and educational outcomes of maltreated and non-maltreated youth: A birth cohort study. Aust N Z J Psychiatry. 2019;53(3):248–55.

9. Strathearn L, Giannotti M, Mills R, Kisely S, Najman J, Abajobir A. Long-term Cognitive, Psychological, and Health Outcomes Associated With Child Abuse and Neglect. Pediatrics. 2020;146(4:e20200438).

10. Gilbert R, Fluke J, O’Donnell M, Gonzalez-Izquierdo A, Brownell M, Gulliver P, et al. Child maltreatment: variation in trends and policies in six developed countries. Lancet. 2012;379:758–72.

11. Rosenthal C M, Parker D M, Thompson L A. Racial Disparities in Child Abuse Medicine. JAMA Pediatrics. 2021;Published online October 18, 2021.

12. Putnam-Hornstein E, Ahn E, Prindle J, Magruder J, Webster DC W. Cumulative rates of child protection involvement and terminations of parental rights in a California birth cohort, 1999–2017. Am J Public Health. 2021;111(6):1157–63.

13. Wildeman C, Emanuel N, Leventhal J M, Putnam-Hornstein E, Waldfogel J, Lee H. The Prevalence of Confirmed Maltreatment Among US Children, 2004 to 2011. JAMA Pediatrics. 2014;168(8):706–13.

14. Kim H, Wildeman C, Jonson-Reid M, Drake B. Lifetime Prevalence of Investigating Child Maltreatment Among US Children. Am J Public Health. 2017;107:274–80.

15. Sinha V, Trocme N, Fallon B, MacLaurin B. Understanding the investigation-stage overrepresentation of First Nations children in the child welfare system: An analysis of the First Nations component of the Canadian Incidence Study of Reported Child Abuse and Neglect 2008. Child Abuse & Neglect. 2013;37(10):821–31.

16. Bilson A, Cant R L, Harries M, Thorpe D H. A Longitudinal Study of Children Reported to the Child Protection Department in Western Australia. The British Journal of Social Work. 2015;45(3):771–91.

17. Rouland B, Vaithianathan R, Wilson D, Putnam-Hornstein E. Ethnic Disparities in Childhood Prevalence of Maltreatment: Evidence From a New Zealand Birth Cohort. Am J Public Health. 2019;109(9):1255–7.

18. Bywaters P, Scourfield J, Webb C, Morris K, Featherstone B, Brady G, et al. Paradoxical evidence on ethnic inequities in child welfare: Towards a research agenda. Children and Youth Services Review. 2019;96:145–54.

19. Bywaters P, Kwhali J, Brady G, Sparks T, Bos E. Out of Sight, Out of Mind: Ethnic Inequalities in Child Protection and Out-of-Home Care Intervention Rates. British Journal of Social Work. 2017;47:1884–902.

20. Kim H, Drake B. Child maltreatment risk as a function of poverty and race/ethnicity in the USA. International Journal of Epidemiology. 2018;47(3):780–7.

21. Putnam-Hornstein E, Needell B. Predictors of child protective service contact between birth and age five: An examination of California’s 2002 birth cohort. Children and Youth Services Review. 2011;33:1337–44.

22. Putnam-Hornstein E, Needell B, King B, Johnson-Motoyama M. Racial and ethnic disparities: A population-based examination of risk factors for involvement with child protective services. Child Abuse & Neglect. 2013;37:33–46.

23. Maguire-Jack K, Lanier P, Johnson-Motoyama M, Welch H, Dineen M. Geographic variation in racial disparities in child maltreatment: The influence of county poverty and population density. Child Abuse & Neglect. 2015;47:1–13.

24. Pham TTL, Berecki-Gisolf J, Clapperton A, O’Brien KS, Liu S, Gibson K. Definitions of Culturally and Linguistically Diverse (CALD): A Literature Review of Epidemiological Research in Australia. Int J Environ Res Public Health. 2021;18.

25. Abdul Rahim R, Pilkington R, D’Onise K, Lynch J. Capturing cultural and linguistic diversity in child health research in Australia. medRxiv. 2020;10.25.20219329.

26. Sawrikar P, Katz I. Recommendations for Improving Cultural Competency When Working with Ethnic Minority Families in Child Protection Systems in Australia. Child Adolesc Soc Work J. 2014;31:25.

27. Kaur J. Cultural Diversity and Child Protection: Australian research review on the needs of culturally and linguistically diverse (CALD) and refugee children and families. Queensland: JK Diversity Consultants, 2012 July 2012. Report No.

28. Lewig K, Arney F, Salveron M. The Working with Refugees Families Project. Australian Centre for Child Protection, University of South Australia, 2009.

29. Sawrikar P, Katz I. ‘Normalizing the Novel’: How Is Culture Addressed in Child Protection Work With Ethnic-Minority Families in Australia? Journal of Social Service Research. 2014;40(1):39–61.

30. Hernan MA, Hsu J, Healy B. A Second Chance to Get Causal Inference Right: A Classification of Data Science Task. Chance. 2019;32(1):42–9.

31. Conroy S, Murray EJ. Let the question determine the methods: descriptive epidemiology done right. British Journal of Cancer 2020;123:1351–2.

32. Pilkington R, Montgomerie A, Grant J, Gialamas A, Malvaso C, Smithers L, et al. An innovative linked data platform to improve the wellbeing of children— the South Australian Early Childhood Data Project. Australia’s welfare 2019 data insights. Australia’s welfare series no. 14. Canberra: AIHW; 2019.

33. SA NT Datalink. Published Papers: SA NT Datalink; 2020 [20/10/2021]. Available from: https://www.santdatalink.org.au/Published_Papers.

34. Procter AM, Pilkington RM, Lynch JW, Smithers LG, Chittleborough CR. Potentially preventable hospitalisations in children: a comparison of definitions. Archives of Disease in Childhood. 2020;105:375–81.

35. Government of South Australia. School Card scheme 2021 [updated 27/01/202110/02/2021]. Available from: https://www.sa.gov.au/topics/education-and-learning/financial-help-scholarships-and-grants/school-card-scheme.

36. Shankar V, Bangdiwala SI. Observer agreement paradoxes in 2×2 tables: comparison of agreement measures. BMC Medical Research Methodology. 2014;14:100.

37. Gwet KL. Computing inter-rater reliability and its variance in the presence of high agreement. British Journal of Mathematical and Statistical Psychology. 2008;61:29–48.

