## Supplementary material for "Child protection contact among children of culturally and linguistically diverse backgrounds: a South Australian linked data study"

### Definition of terms that describe child protection contact in South Australia^1^

**Notification:** A report (allegation) to the agency concerning suspected child abuse or neglect.

**Screened-in:** A notification assessed as meeting the threshold for intervention and therefore requiring a response by the agency.

**Investigation:** A determination of whether an incident of child abuse or neglect has occurred, and the circumstances of its occurrence.

**Substantiation:** A professional judgement as to whether abuse or neglect has occurred.

**Out-of-home care (OOHC):** System of caring for a child who is removed from their family of origin. Includes (but is not limited to) home-based care, emergency care and residential care.

### Figure S1

Figure S1A and B show reports have increased among CALD and non-CALD groups from 2009-15. Among the oldest cohort who were in reception in 2009, 1 in 5 non-CALD children and 1 in 10 CALD children were reported by age 4 years. This increased to 1 in 4 non-CALD children and 1 in 7 CALD children in the 2015 school reception cohort. Cumulative incidence by age 7 in non-CALD children increased from 30.7% in the 2009 cohort to 36.1% in the 2015 cohort. Cumulative incidence of reports by age 7 in CALD children increased from 17.8% in the 2009 cohort to 21.7% in the 2015 cohort.

A: CALD

Cumulative incidence (%)

| Age (years) | 2009 | 2010 | 2011 | 2012 | 2013 | 2014 | 2015 |
| --- | --- | --- | --- | --- | --- | --- | --- |
| 0 | 2.8 | 3 | 3 | 3.4 | 3.6 | 3.6 | 5 |
| 1 | 4.2 | 4.8 | 6.4 | 5.8 | 6.5 | 5.8 | 7.3 |
| 2 | 6 | 7.3 | 9.1 | 8.3 | 9.5 | 8.3 | 9.5 |
| 3 | 7.9 | 9.1 | 11.8 | 10.6 | 11.9 | 10.7 | 11.9 |
| 4 | 10.5 | 12 | 14.4 | 13 | 14.5 | 12.5 | 14.7 |
| 5 | 13.5 | 14.7 | 18.9 | 15.8 | 17.6 | 16.7 | 17.3 |
| 6 | 15.5 | 17.2 | 21.4 | 18.5 | 20 | 20.3 | 19.2 |
| 7 | 17.8 | 19.3 | 24.2 | 21.5 | 22.3 | 22.2 | 21.7 |

B: Non-CALD

Cumulative incidence (%)

| Age (years) | 2009 | 2010 | 2011 | 2012 | 2013 | 2014 | 2015 |
| --- | --- | --- | --- | --- | --- | --- | --- |
| 0 | 5.7 | 6.5 | 6.3 | 7.2 | 8.4 | 8.7 | 8.9 |
| 1 | 9.7 | 10.9 | 11.5 | 12.3 | 13.9 | 13.7 | 14.1 |
| 2 | 12.9 | 15.1 | 15.4 | 17.2 | 17.8 | 17.5 | 17.9 |
| 3 | 16.8 | 19.3 | 19 | 21.2 | 21.3 | 21.2 | 21.5 |
| 4 | 20.4 | 23.3 | 22.5 | 25.5 | 24.8 | 25.7 | 25.3 |
| 5 | 24.2 | 27 | 26.3 | 29.4 | 28.6 | 29.8 | 29.6 |
| 6 | 27.7 | 30.4 | 29.9 | 33.4 | 32.9 | 33.9 | 33.3 |
| 7 | 30.7 | 33.8 | 33.2 | 37.2 | 37 | 37.9 | 36.1 |

Figure S1: Cumulative incidence of child protection reports by age and year commenced Reception, 2009 – 2015 (A): CALD (B): non-CALD

### **Agreement between two measures of CALD**

The AC_1_ is computed as follows:

$$\hat{AC_{1}} =\frac{P_{0} - P_{e}^{G}}{1 - P_{e}^{G}}$$

where P_0_ = the proportion of units where the two definitions agreed (concordance),

P_0_ = (x_11_ + x_22_)/N

P_e_ = the proportion of units where agreement is expected by chance ^2^ (expected concordance)

P_e_^G^ = 2*((f_1_/N + g_1_/N)/2) * [1 – ((f_1_/N + g_1_/N)/2)] ^3^

Where x_11_ and x_22_, are the concordant cells and x_12_ and x_21_ are the discordant cells in a classification matrix represented by a 2x2 table. Similar to Cohen’s kappa an AC_1_ of 1 indicates perfect agreement between two measures.

Table S1 shows according to the preferred definition, the proportion of CALD and non-CALD children was 16.3% and 83.7% respectively. Using the alternative definition, CALD children comprised 13.9% of the population. As the distributions were skewed we used the AC_1_ instead of Cohen’s kappa given the limitation of this measure when there is imbalance in the distribution of outcome classes. ^3, 4^ The AC_1_ is similar to Cohen’s kappa in its formulation but only differs in how the expected probabilities are derived.^3, 4^ The AC_1_ value was 0.92, demonstrating a high level of agreement between the two definitions.

Table S1: Number of children classified as CALD and non-CALD by the preferred and alternative definitions of CALD (N = 34,217)

|  | | **Preferred definition** | | **Total** |
| --- | --- | --- | --- | --- |
|  |  | **CALD** (1) | **Non-CALD**  (2) |  |
| **Alternative definition** | **CALD**  (1) | 4175 (X_11_) | 575 (X_12_) | 4750 ($g_{1}$) |
|  | **Non-CALD**  (2) | 1421 (X_21_) | 28 100 (X_22_) | 29 521 ($g_{2}$) |
|  | **Total** | 5596 ($f_{1}$) | 28 675 ($f_{2}$) | 34 271 ($N$) |

### **Child protection contact under variations to the study population**

In the first sensitivity analysis (Table S2) that included both independent and government school children, results showed that by age 7, 9.5% of CALD compared to 14.5% of non-CALD children had been screened in. By age 7, 0.3% of CALD children had experienced OOHC compared to 1.7% of non-CALD (RD 1.4% (95% CI 1.2%-1.6%)). In sensitivity analysis two (Table S3), 11.4% of CALD children were screened in by age 7 compared to 18.3% of non-CALD children with risk difference of 6.9% (95% CI 5.8%-8.0%). By age 7, 0.6% of CALD children had experienced OOHC compared to 2.1% of non-CALD children (RD 1.5%, 95% CI 1.2%-1.8%). In sensitivity analysis three (Table S4) where Aboriginal and Torres Strait Islander children were excluded, the overall pattern of contact remained the same. By age 7, the proportion of CALD screened in was 11.1% compared to 16.3% of non-CALD children (RD 5.1%, 95% CI 4.4%–5.8%). The proportion of CALD and non-CALD children that had experienced OOHC by age 7 were 0.6% and 1.6% respectively, corresponding to a risk difference of 1.0% (95% CI 0.8% - 1.2%). In sensitivity analysis four which expanded the study population to include migrant children who arrived in SA after birth, the patterns of contact between the two groups were preserved as shown in Tables S5-S7.

Table S2: Child protection contact by ages 4 and 7 years for children born in SA and sat the AEDC in SA in years 2009, 2012 and 2015, followed to the end of 2017 (n=46 759)

| Child protection contact | Non-CALD  n = 41 921 | CALD  n = 4838 | Total  N = 46 759 | Risk difference (95% CI)  (Non-CALD – CALD) | Risk ratio (95% CI)  (Non-CALD/ CALD) |
| --- | --- | --- | --- | --- | --- |
|  | % with contact | % with contact | N (%) |  |  |
| **By age 4 (prior to school years)** | | | | | |
| Reported | 15.5 | 10.0 | 6996 (15.0) | 5.6 (4.7, 6.5) | 1.56 (1.43, 1.70) |
| Screened in | 11.1 | 6.5 | 4964 (10.6) | 4.6 (3.8, 5.4) | 1.71 (1.53, 1.91) |
| Investigated | 6.0 | 2.5 | 2630 (5.6) | 3.5 (3.0, 4.0) | 2.41 (2.01, 2.89) |
| Substantiated | 3.0 | 1.0 | 1323 (2.8) | 2.0 (1.7, 2.3) | 2.94 (2.22, 3.89) |
| OOHC | 1.3 | 0.3 | 563 (1.2) | 1.0 (0.9, 1.2) | 4.89 (2.82, 8.46) |
| **By age 7 (including first 3 years of school)** | | | | | |
| Reported | 21.2 | 15.6 | 9625 (20.6) | 5.5 (4.4, 6.6) | 1.35 (1.26, 1.43) |
| Screened in | 14.5 | 9.5 | 6540 (14.0) | 5.0 (4.1, 5.9) | 1.53 (1.39, 1.67) |
| Investigated | 7.4 | 3.2 | 3254 (7.0) | 4.2 (3.7, 4.8) | 2.34 (1.99, 2.74) |
| Substantiated | 4.0 | 1.3 | 1746 (3.7) | 2.8 (2.4, 3.1) | 3.19 (2.47, 4.11) |
| OOHC | 1.7 | 0.3 | 710 (1.5) | 1.4 (1.2, 1.6) | 5.74 (3.38, 9.73) |

Table S3: Child protection contact by ages 4 and 7 years for children born in SA and sat the AEDC in SA government school in years 2009, 2012 and 2015, followed to the end of 2017 (n=29 172)

| Child protection contact | Non-CALD  n = 25 180 | CALD  n = 3992 | Total  N = 29 172 | Risk difference (95% CI)  (Non-CALD – CALD) | Risk ratio (95% CI)  (Non-CALD/ CALD) |
| --- | --- | --- | --- | --- | --- |
|  | % with contact | % with contact | N (%) |  |  |
| **By age 4 (prior to school years)** | | | | | |
| Reported | 19.7 | 12.3 | 5444 (18.7) | 7.4 (6.3, 8.5) | 1.60 (1.47, 1.75) |
| Screened in | 14.2 | 8.0 | 3902 (13.4) | 6.2 (5.3, 7.2) | 1.77 (1.59, 1.98) |
| Investigated | 7.8 | 3.4 | 2086 (7.2) | 4.4 (3.7, 5.0) | 2.31 (1.94, 2.74) |
| Substantiated | 4.0 | 1.5 | 1059 (3.6) | 2.4 (2.0, 2.9) | 2.59 (2.01, 3.35) |
| OOHC | 1.6 | 0.6 | 428 (1.5) | 1.0 (0.8, 1.3) | 2.79 (1.84, 4.24) |
| **By age 7 (including first 3 years of school)** | | | | | |
| Reported | 26.0 | 18.5 | 7294 (25.0) | 7.5 (6.2, 8.8) | 1.40 (1.31, 1.50) |
| Screened in | 18.3 | 11.4 | 5056 (17.3) | 6.9 (5.8, 8.0) | 1.60 (1.46, 1.75) |
| Investigated | 9.6 | 4.2 | 2573 (8.8) | 5.4 (4.7, 6.1) | 2.28 (1.96, 2.66) |
| Substantiated | 5.3 | 1.8 | 1409 (4.8) | 3.4 (2.9, 3.9) | 2.86 (2.27, 3.61) |
| OOHC | 2.1 | 0.6 | 557 (1.9) | 1.5 (1.2, 1.8) | 3.52 (2.34, 5.29) |

Table S4: Child protection contact by ages 4 and 7 years for non-Indigenous children born in SA and attended reception in government schools between years 2009 and 2015, followed to the end of 2017 (n=72 394)

| Child protection contact | Non-CALD  n = 63 170 | CALD  n = 9224 | Total  N = 72 394 | Risk difference (95% CI)  (Non-CALD – CALD) | Risk ratio (95% CI)  (Non-CALD/ CALD) |
| --- | --- | --- | --- | --- | --- |
|  | % with contact | % with contact | N (%) |  |  |
| **By age 4 (prior to school years)** | | | | | |
| Reported | 17.3 | 11.9 | 12 043 (16.6) | 5.4 (4.6, 6.1) | 1.45 (1.37, 1.54) |
| Screened in | 12.3 | 8.0 | 8533 (11.8) | 4.3 (3.7, 4.9) | 1.53 (1.43, 1.65) |
| Investigated | 6.4 | 3.4 | 4373 (6.0) | 3.0 (2.6, 3.4) | 1.89 (1.69, 2.12) |
| Substantiated | 3.2 | 1.5 | 2154 (3.0) | 1.7 (1.4, 2.0) | 2.18 (1.84, 2.30) |
| OOHC | 1.2 | 0.5 | 817 (1.1) | 0.7 (0.5, 0.9) | 2.29 (1.72, 3.05) |
| **By age 7 (including first 3 years of school)** | | | | | |
| Reported | 23.8 | 17.9 | 16677 (23.0) | 5.9 (5.0, 6.7) | 1.33 (1.27, 1.39) |
| Screened in | 16.3 | 11.2 | 11343 (15.7) | 5.1 (4.4, 5.8) | 1.46 (1.37, 1.55) |
| Investigated | 8.1 | 4.2 | 5492 (7.6) | 3.9 (3.4, 4.3) | 1.92 (1.74, 2.12) |
| Substantiated | 4.4 | 1.9 | 2929 (4.1) | 2.5 (2.2, 2.8) | 2.34 (2.01, 2.73) |
| OOHC | 1.6 | 0.6 | 1071 (1.5) | 1.0 (0.8, 1.2) | 2.60 (1.99, 3.39) |

Table S5: Child protection contact by ages 4 and 7 years for all children who attended reception in government schools between years 2009 and 2015, followed to the end of 2017 (n=91 395)

| Child protection contact | Non-CALD  n = 76 123 | CALD  n = 15 272 | Total  N = 91 395 | Risk difference (95% CI)  (Non-CALD – CALD) | Risk ratio (95% CI)  (Non-CALD/ CALD) |
| --- | --- | --- | --- | --- | --- |
|  | % with contact | % with contact | N (%) |  |  |
| **By age 4 (prior to school years)** | | | | | |
| Reported | 19.0 | 9.1 | 15 850 (17.3) | 9.8 (9.3, 10.4) | 2.08 (1.97, 2.19) |
| Screened in | 14.0 | 6.1 | 11 581 (12.7) | 7.9 (7.5, 8.4) | 2.31 (2.16, 2.46) |
| Investigated | 7.7 | 2.4 | 6214 (6.8) | 5.3 (5.0, 5.6) | 3.18 (2.86, 3.53) |
| Substantiated | 4.0 | 1.0 | 3198 (3.5) | 3.0 (2.7, 3.2) | 3.83 (3.27, 4.49) |
| OOHC | 1.6 | 0.4 | 1254 (1.4) | 1.1 (0.9, 1.2) | 5.87 (3.67, 9.37) |
| **By age 7 (including first 3 years of school)** | | | | | |
| Reported | 25.9 | 14.8 | 21 984 (24.1) | 11.2 (10.5, 11.8) | 1.76 (1.69, 1.83) |
| Screened in | 18.4 | 9.0 | 15 374 (16.8) | 9.3 (8.8, 9.9) | 2.03 (1.93, 2.14) |
| Investigated | 9.8 | 3.2 | 7914 (8.7) | 6.6 (6.2, 6.9) | 3.07 (2.81, 3.36) |
| Substantiated | 5.5 | 1.5 | 4407 (4.8) | 4.0 (3.8, 4.3) | 3.69 (3.24, 4.22) |
| OOHC | 2.1 | 0.4 | 1694 (1.8) | 1.4 (1.2, 1.5) | 5.92 (3.91, 8.96) |

Table S6: Child protection contact by ages 4 and 7 years for all children who sat the AEDC in SA in government and independent schools in years 2009, 2012 and 2015, followed to the end of 2017 (n=55 232)

| Child protection contact | Non-CALD  n = 46 931 | CALD  n = 8301 | Total  N = 55 232 | Risk difference (95% CI)  (Non-CALD – CALD) | Risk ratio (95% CI)  (Non-CALD/ CALD) |
| --- | --- | --- | --- | --- | --- |
|  | % with contact | % with contact | N (%) |  |  |
| **By age 4 (prior to school years)** | | | | | |
| Reported | 14.9 | 7.6 | 7647 (13.8) | 7.3 (6.7, 8.0) | 1.96 (1.81, 2.12) |
| Screened in | 10.6 | 4.9 | 5405 (9.8) | 5.7 (5.2, 6.3) | 2.16 (1.96, 2.38) |
| Investigated | 5.7 | 1.8 | 2841 (5.1) | 3.9 (3.6, 4.3) | 3.20 (2.71, 3.76) |
| Substantiated | 2.9 | 0.8 | 1447 (2.6) | 2.1 (1.9, 2.4) | 3.64 (2.85, 4.65) |
| OOHC | 1.3 | 0.2 | 615 (1.1) | 1.2 (1.0, 1.4) | 3.88 (2.60, 5.80) |
| **By age 7 (including first 3 years of school)** | | | | | |
| Reported | 20.6 | 13.1 | 10 783 (19.5) | 7.5 (6.7, 8.3) | 1.57 (1.48, 1.67) |
| Screened in | 14.1 | 7.8 | 7281 (13.2) | 6.3 (5.6, 6.9) | 1.80 (1.67, 1.95) |
| Investigated | 7.2 | 2.4 | 3590 (6.5) | 4.8 (4.4, 5.2) | 2.97 (2.58, 3.41) |
| Substantiated | 4.0 | 1.1 | 1945 (3.5) | 2.9 (2.6, 3.2) | 3.78 (3.05, 4.68) |
| OOHC | 1.6 | 0.3 | 793 (1.4) | 1.7 (1.4, 1.9) | 4.67 (3.20, 6.81) |

Table S7: Child protection contact by ages 4 and 7 years for all children who sat the AEDC in SA government schools in years 2009, 2012 and 2015, followed to the end of 2017 (n=34 271)

| Child protection contact | Non-CALD  n = 28 100 | CALD  n = 6171 | Total  N = 34 271 | Risk difference (95% CI)  (Non-CALD – CALD) | Risk ratio (95% CI)  (Non-CALD/ CALD) |
| --- | --- | --- | --- | --- | --- |
|  | % with contact | % with contact | N (%) |  |  |
| **By age 4 (prior to school years)** | | | | | |
| Reported | 19.1 | 9.9 | 5976 (17.4) | 9.1 (8.3, 10.0) | 1.92 (1.77, 2.08) |
| Screened in | 13.8 | 6.5 | 4270 (12.5) | 7.3 (6.5, 8.0) | 2.11 (1.91, 2.33) |
| Investigated | 7.5 | 2.5 | 2262 (6.6) | 5.0 (4.5, 5.5) | 2.96 (2.53, 3.48) |
| Substantiated | 3.9 | 1.2 | 1164 (3.4) | 2.7 (2.3, 3.0) | 3.23 (2.56, 4.09) |
| OOHC | 1.6 | 0.4 | 467 (1.4) | 1.2 (1.1, 1.4) | 4.46 (3.40, 5.85) |
| **By age 7 (including first 3 years of school)** | | | | | |
| Reported | 25.7 | 15.9 | 8191 (23.9) | 9.7 (8.7, 10.8) | 1.61 (1.52, 1.71) |
| Screened in | 18.0 | 9.6 | 5643 (16.5) | 8.4 (7.5, 9.2) | 1.87 (1.73, 2.03) |
| Investigated | 9.4 | 3.3 | 2849 (8.3) | 6.1 (5.6, 6.7) | 2.86 (2.49, 3.29) |
| Substantiated | 5.3 | 1.5 | 1573 (4.6) | 3.7 (3.3, 4.1) | 3.42 (2.78, 4.20) |
| OOHC | 2.1 | 0.4 | 623 (1.8) | 1.7 (1.6, 1.8) | 4.87 (3.82, 6.22) |

ReRe

ReferencesRe

ReRe

1.	Child Protection Systems Royal Commission. The life they deserve: Child Protection Systems Royal Commission Report. 2016.

2.	Cohen J. Coefficient of Agreement for Nominal Scales. Educational and Psychological Measurement. 1960;20(1):37-46.

3.	Shankar V, Bangdiwala SI. Observer agreement paradoxes in 2x2 tables: comparison of agreement measures. BMC Medical Research Methodology. 2014;14:100.

4.	Gwet KL. Computing inter-rater reliability and its variance in the presence of high agreement. British Journal of Mathematical and Statistical Psychology. 2008;61:29-48.
